# Disparities in SARS-CoV-2 Vaccination-to-Infection Risk, Massachusetts 2020-2021

**DOI:** 10.1101/2021.04.14.21255467

**Authors:** Scott Dryden-Peterson, Gustavo E. Velásquez, Thomas J. Stopka, Sonya Davey, Rajesh T. Gandhi, Shahin Lockman, Bisola O. Ojikutu

## Abstract

**Background:** Effective vaccine-based containment strategies for SARS-CoV-2 require equitable coverage of communities at greatest risk of infection. We sought to examine the alignment of vaccination and SARS-CoV-2 risk in Massachusetts to inform public health response.

**Methods:** We aggregated cumulative SARS-CoV-2 testing and vaccination data from the Massachusetts Department of Public Health and the Boston Public Health Commission from January 29, 2020 to April 9, 2021. We used two approaches to assess vaccination equity: vaccination-to-infection risk (VIR) ratio and Lorenz curves. The VIR ratio was calculated for each community as the quotient of the number of fully vaccinated individuals divided by the cumulative number of confirmed SARS-CoV-2 infections. Lorenz curves were used to describe vaccination relative to COVID-19 burden. A multivariable Poisson model was used to assess predictors of VIR ratio.

**Results:** A total of 607,120 (8.9%) SARS-CoV-2 infections were confirmed in Massachusetts residents and 1,485,266 (21.8%) residents were fully vaccinated. Communities with increased socioeconomic vulnerability had lower VIR ratios indicating less equitable vaccination relative to infection risk. In multivariable analysis, decreased vaccination relative to infection risk was independently associated with increasing socioeconomic vulnerability (aRR 0.82 per quartile increase, 95% CI 0.77 to 0.87) and with greater than 20% of the community identified as Black and/or Latinx (aRR 0.67, 95% CI 0.56 to 0.81). Improved community vaccine delivery was associated with higher community proportion of residents aged 65 or older (aRR 1.23 per 5% increase in proportion, 95% CI 1.15 to 1.31). Lorenz curves indicated considerable inequity (Gini 0.46 between communities). An estimated 330,000 full vaccination courses would need to be diverted to under-vaccinated communities to achieve equity.

**Conclusion:** In conclusion, disparities in vaccine delivery highlight ongoing inequities in our approach to COVID-19 and imperil efforts to control the pandemic.

## Introduction

Effective vaccine-based containment strategies for SARS-CoV-2 require equitable coverage of communities at greatest risk of infection.^1^ However, significant disparities in vaccination rates have been observed^2^. Adapting a concept from HIV prevention efforts^3^, we examined the alignment of vaccination and SARS-CoV-2 risk in Massachusetts, creating and applying a vaccination-to-infection risk ratio.

## Methods

We aggregated cumulative SARS-CoV-2 testing and vaccination data from the Massachusetts Department of Public Health (MDPH) and the Boston Public Health Commission from January 29, 2020 to April 9, 2021. We limited analysis to 291 communities (276 cities and towns; 15 Boston neighborhoods) with a population over 3,000 residents (total population 6,795,000).

We considered each community’s cumulative incidence of confirmed SARS-CoV-2 infections to be the best available indicator of future infection risk. We used two approaches to assess vaccination equity: vaccination-to-infection risk (VIR) ratio and Lorenz curves. The VIR ratio was calculated for each community as the quotient of the number of fully vaccinated individuals divided by the cumulative number of confirmed SARS-CoV-2 infections. Communities with VIR ratios below the state-wide mean have lower vaccination coverage relative to their infection risk. Lorenz curves, which assess equity in resource distribution^4^, were used to describe vaccination relative to COVID-19 burden and calculate summaries of inequity (Gini index) and magnitude of vaccine re-allocation required to achieve equity (Hoover index).

We utilized population estimates from MDPH and the American Community Survey (2014-2018) to determine community age, race, and ethnic composition. Socioeconomic vulnerability was estimated using the Socioeconomic Status domain of the Social Vulnerability Index.^5^ We fit a Poisson model utilizing robust sandwich estimators to assess associations between community VIR ratios and a priori selected predictors: proportion aged 65 years or older (an early vaccination eligibility criterion), proportion identified as Black and/or Latinx (<20% or ≥20%), quartile of socioeconomic vulnerability, and community population (<50,000 or ≥50,000 residents). We adhered to STROBE guidelines; this work was designated as exempt by the Mass General Brigham IRB.

## Results

As of April 9, 2021, 607,120 (8.9%) SARS-CoV-2 infections had been confirmed in Massachusetts residents and 1,485,266 (21.8%) residents were fully vaccinated. Cumulative incidence of confirmed SARS-CoV-2 infection (minimum: 1.9%, maximum: 22.7%) and complete vaccination (minimum: 10.1%, maximum: 53%) varied considerably between communities. Communities with increased socioeconomic vulnerability had lower VIR ratios indicating less equitable vaccination relative to infection risk (**Figure 1**).

**Figure 1.**
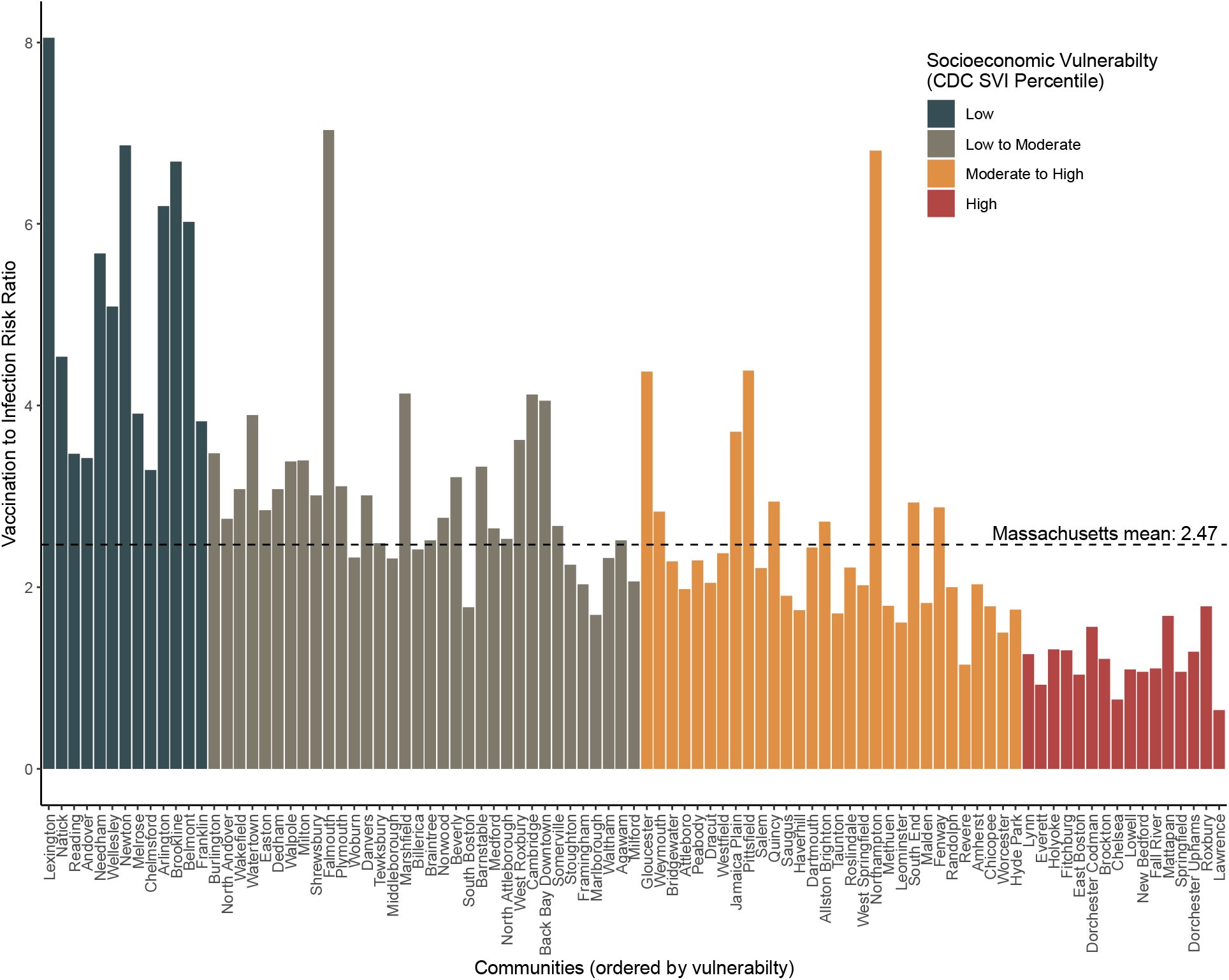
The vaccination-to-infection risk (VIR) ratio among Massachusetts communities with a population greater than 25,000 residents, ordered by socioeconomic vulnerability. The VIR ratio is calculated as the cumulative number of fully vaccinated individuals divided by the cumulative number of confirmed SARS-CoV-2 infections in each community reported from January 29, 2020 through April 9, 2021.

In multivariable analysis, decreased vaccination relative to infection risk was independently associated with increasing socioeconomic vulnerability (adjusted relative risk [aRR] 0.82 per quartile increase, 95% CI 0.77 to 0.87, p<0.001) and with greater than 20% of the community identified as Black and/or Latinx (aRR 0.67, 95% CI 0.56 to 0.81, p<0.001). Improved community vaccine delivery was associated with higher community proportion of residents aged 65 or older (aRR 1.23 per 5% increase in proportion, 95% CI 1.15 to 1.31, p<0.001). Community population size was not associated with the VIR ratio (p=0.65).

Lorenz curves indicated considerable inequity, with an estimated Gini coefficient (1, perfect equity; 0, perfect inequity) of 0.46 between communities and 0.51 by individuals statewide (**Figure 2**). An estimated 330,000 full vaccination courses would need to be diverted to under-vaccinated communities to achieve equity.

**Figure 2.**
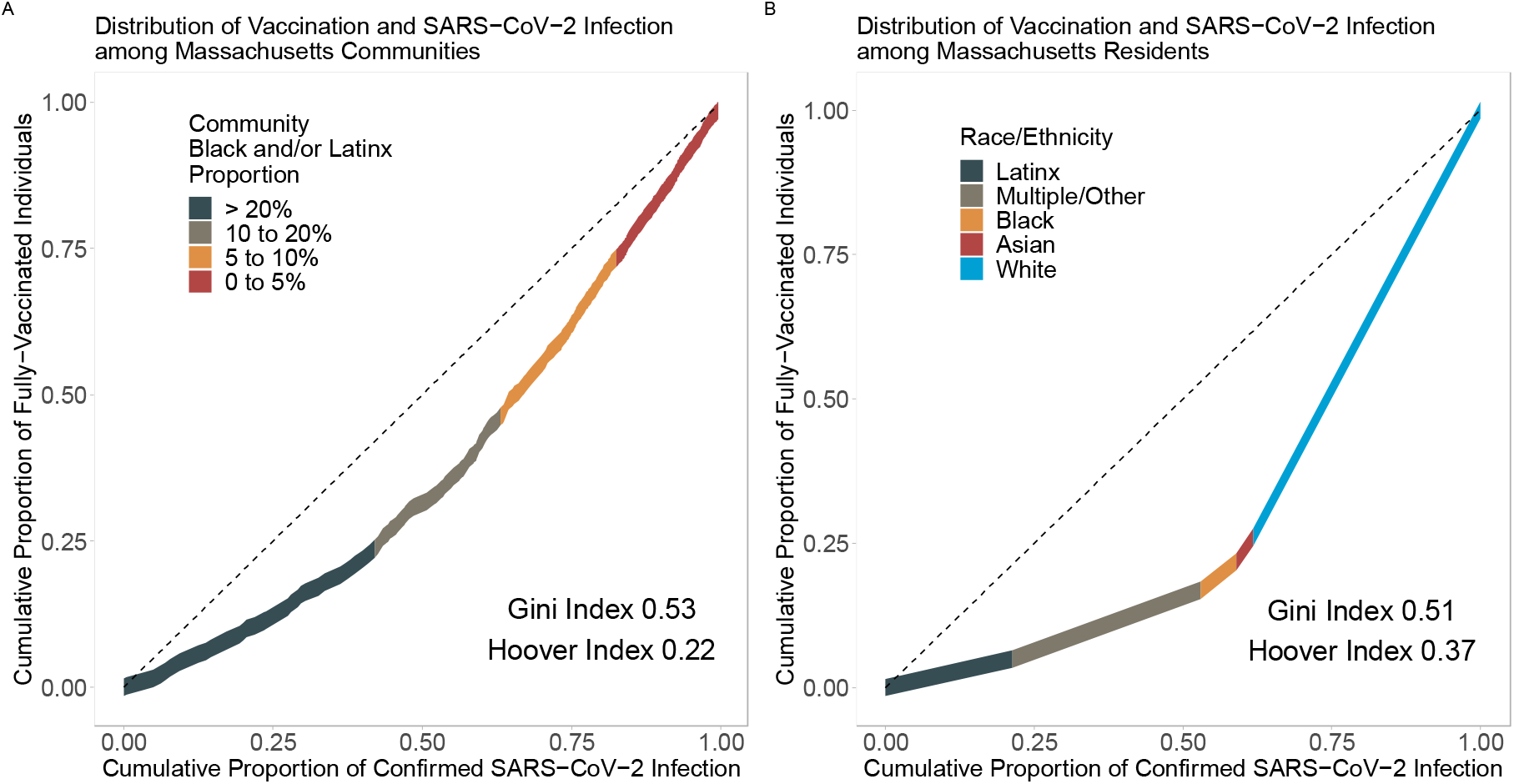
Lorenz curves of cumulative proportion of individuals who have been fully vaccinated against SARS-CoV-2 versus the cumulative proportion of confirmed COVID-19 infections by communities (A) and individual residents (B) in Massachusetts, grouped by measures of racial and ethnic composition. Non-White residents account for 62% of confirmed infections and only 26% of full vaccinations. Cases and vaccinations reported from January 29, 2020 through April 9, 2021 are included.

## Discussion

These analyses of SARS-CoV-2 vaccination indicate lower vaccine delivery relative to infection risk in socioeconomically vulnerable communities and those with larger proportions of Black and Latinx populations. The disparities reported here are greater than those previously reported using county-level data^2^, and emphasize the importance of monitoring and targeting smaller geographic units to guide equitable distribution of vaccines.^6^

In conclusion, disparities in vaccine delivery highlight ongoing inequities in our approach to COVID-19 and imperil efforts to control the pandemic.

## Data Availability

All data and analytic code to be made available

https://github.com/sldrydenpeterson/MA-SARSCoV2-Testing-Alignment.git

## Acknowledgements

The work was made possible with help from the Harvard University Center for AIDS Research (CFAR), a funded program of the National Institutes of Health (P30 AI060354), the National Institute of Allergy and Infectious Diseases (K08 AI141740, K24 AI131928, UM1 AI069412), the Dr. Lynne Reid/Drs. Eleanor and Miles Shore Fellowship at Harvard Medical School, and the Burke Global Health Fellowship at the Harvard Global Health Institute. The contents of this manuscript are solely the responsibility of the authors and do not necessarily represent the official views of the National Institutes of Health or the institutions with which the authors are affiliated. The funding source had no role in the design and conduct of the study; collection, management, analysis, and interpretation of the data; preparation, review, or approval of the manuscript; and decision to submit the manuscript for publication. Dr. Dryden-Peterson had full access to all the data in the study and takes responsibility for the integrity of the data and the accuracy of the data analysis.

